# Regional variation and epidemiological insights in malaria underestimation in Cameroon

**DOI:** 10.1101/2023.11.06.23298167

**Authors:** Sarafa A. Iyaniwura, Qing Han, Ngem Bede Yong, Ghislain Rutayisire, Agnes Adom-Konadu, Okwen Patrick Mbah, David Poumo Tchouassi, Kingsley Badu, Jude D. Kong

**Affiliations:** Theoretical Biology and Biophysics, Theoretical Division, Los Alamos National, Los Alamos, NM 87545, USA; Laboratory for Industrial and Applied Mathematics (LIAM), Department of Mathematics and Statistics, York University, Toronto, M3J 1P3, Ontario, Canada; Africa-Canada Artificial Intelligence and Data Innovation Consortium (ACADIC), York University, Toronto, M3J 1P3, Ontario, Canada; Department of Mathematics and Statistics, York University, Toronto, M3J 1P3, Ontario, Canada; Department of Mathematics, University of Cape Coast, Cape Coast, Central Region, Ghana; Department of Public Health, Regional Delegation of Public Health North West Region, Bamenda, Cameroon; Effective Basic Services Africa (eBASE), Research and Development, Bamenda, North West Region, Cameroon; International Centre of Insect Physiology and Ecology, Nairobi, Kenya; Department of Theoretical and Applied Biology, Kwame Nkrumah University of Science and Technology, University Post Office, Kumasi AK 039, Ghana

## Abstract

Malaria, caused by *Plasmodium* parasites and transmitted by female *Anopheles* mosquitoes, is most common in tropical regions, especially in Sub-Saharan Africa. Despite significant global effort to control and eradicate the disease, many cases and deaths are still reported yearly. These efforts are hindered by several factors, including the severe underestimation of cases and deaths, especially in Africa, making it difficult to assess the disease burden accurately. We used a mathematical model of malaria, incorporating the underestimation of cases and seasonality in mosquito biting rate, to study the disease dynamics in Cameroon. Using a Bayesian inference framework, we calibrated our model to the monthly reported malaria cases in ten regions of Cameroon from January 2019 to December 2021 to quantify the underestimation of cases and estimate other important epidemiological parameters. We performed Hierarchical Clustering on Principal Components analysis to understand regional disparities, looking at underestimation rates, population sizes, healthcare personnel, and healthcare facilities per 1,000 people. We found varying levels of underestimation of cases across regions, with the East region having the lowest underestimation (14%) and the Northwest region with the highest (70%). The mosquito biting rate peaks once every year in most of the regions, except in the Northwest region where it peaks every 6.02 months and in Littoral every 15 months. We estimated a median mosquito biting rate of over five bites per day for most of the regions with Littoral having the highest (9.86 bites/day). Two regions have rates below five bites per day: Adamawa (4.78 bites/day) and East (4.64 bites/day). The notably low estimation of malaria cases in Cameroon underscore the pressing requirement to bolster reporting and surveillance systems. Regions in Cameroon display a range of unique features, which may contribute to the differing levels of malaria underestimation. These distinctions should be considered when evaluating the efficacy of community-based interventions.

**Author summary:**

i. We used a deterministic mathematical model of malaria that incorporated the underestimation of cases and seasonality in the biting rate of mosquitoes to retroactively study the dynamics of the disease in Cameroon from January 2019 to December 2021.
ii. We found varying levels of underestimation of malaria cases across regions in Cameroon, with the East region having 14% underestimation and the Northwest region having 70%.
iii. We found consistent malaria-induced death rates and natural immunity duration across Cameroon. We estimated that the mosquito biting rate for the Northwest region oscillated with a period of 6.02 months, while those of the remaining regions had a period of 12 months or more. Most regions had median mosquito biting rates exceeding five bites per day, with the Littoral having the highest (9.86 bites/day). In comparison, two regions had rates below five bites per day: Adamawa (4.78 bites/day) and East (4.64 bites/day).
iv. We clustered the ten regions into four major groups using the case underestimation rate, population size, total healthcare human resources per 1,000, and total healthcare facilities per 1,000.

## Introduction

Malaria is a mosquito-borne disease caused by *plasmodium* parasites and transmitted by female *Anopheles* mosquitoes. It is common in sub-Saharan Africa and other tropical regions of the world [1–3]. There are 123 known species of the genus *Plasmodium*. However, only five of them are known to cause infections in human: *P. falciparum, P. malariae, P. ovale, P. vivax*, and *P. knowlesi* [7–11]. The most prevalent and dangerous malaria parasite is *Plasmodium falciparum* [12]. It is responsible for a significant portion of malaria-related fatalities [7, 8, 13]. *Plasmodium* parasites are mainly spread through mosquito bites, but can also be spread from a malaria-infected pregnant individual to their fetus through the placenta, and through blood transfusion [14]. Individuals infected with malaria may have symptoms like fever, rigors, and chills, in the case of uncomplicated malaria [15]. However, severe malaria mostly in children under five may be presented as fever, impaired consciousness, severe anaemia, respiratory distress, convulsions, and hypoglycemia, among other symptoms [8, 16]. Symptoms usually start 10 to 15 days after the initial mosquito bite, however for infections involving some strains of *plasmodium vivax*, symptoms could delay for 8 to 10 months or longer [17].

About half of the world’s population, the majority of whom reside in Africa, are susceptible to malaria and dealing with its economic hardships. According to the WHO estimates, there were 247 million malaria cases and 619, 000 deaths in 2021, where 234 million (∼ 95%) of the cases and 593, 000 (∼ 96%) of the deaths were in Africa [12]. In addition, ∼79% of the deaths were in children under five years [12]. Despite recent advancements in the fight against malaria, the disease is still widespread with over 2 million reported cases in Cameroon yearly, posing a serious public health concern [18–22]. It is identified as the third leading cause of death in healthcare centers in Cameroon, accounting for 30% of the mortality [23].

The burden of malaria differs geographically depending on the local malaria transmission intensity [24]. Underestimation of cases can be in terms of under-ascertainment (when not all patients seek healthcare), under-diagnosis (when cases seek healthcare but are not properly diagnosed) and/or under-reporting or under-notification (when diagnosed cases are not adequately reported and notified) [25]. The burden of asymptomatic malaria, i.e., infection with *P. falciparum* in the absence of overt clinical symptoms, is huge [3]. Individuals may harbor malaria parasites but due to the absence of symptoms will not report to any facility for diagnosis or treatment and thus will not be counted. In addition, some individuals may self-treat, either with local herbs or through medication from undocumented sources. These may lead to the under-ascertainment of cases. Furthermore, infected individuals may visit hospitals but their cases go undetected due to low parasite load and/or the sensitivity of the testing device [26]. Recently, Afriyie et al. [26] found that up to 40% of individuals diagnosed as negative in the lab by microscopy and rapid diagnostic tests (RDT) in the Ashanti Region of Ghana, were identified as positive under research settings when tested with molecular techniques with higher sensitivity. Such individuals with undetected infection bring concern to malaria elimination in Africa, as they present as a reservoir for mosquitoes to pick up infections and spread to others.

One of the earliest mathematical models used to study malaria transmission is the “Ross-Macdonald model” developed in 1915 [27]. Over the years, this model has led to the development of other deterministic mathematical models used to gain insight into the transmission dynamics of malaria [7, 28, 29, 29–31]. Of recent, Ndamuzi et al. [7] proposed a deterministic model for studying the dynamics of malaria parasite in mosquito and human populations. They demonstrated the need for effective control measures to decrease the number of mosquito bites per individual per unit time and the population of mosquitoes, among other factors to adequately control the spread of malaria. Another malaria dynamics model with age-structure in mosquito population was developed by Bakary et al. [10]. The model divides the human population into non-immune and semi-immune, where the non-immune are the most vulnerable, while the semi-immune are the least vulnerable individuals. They analyzed the model mathematically and studied the stability properties of its steady-states. The model of Collins et al. [32] was used to study the dynamics of malaria in Nigeria. The model incorporates the treatment of malaria, drug resistance, and the usage of mosquito nets as a preventive measure. Their finding suggests that unless better control efforts are directed at the dominant resistant strain of the disease, treatment is improved, and mosquito nets are used widely, malaria is likely to remain endemic in Nigeria.

The underestimation of malaria cases has been reported in some regions around the world [46–49, 51, 52]. In an India forest community, Chourasia et al. [46] estimated that malaria cases maybe have been underestimated by up to 20% and 22.8% in 2013 and 2014, respectively, due to asymptomatic cases. A cross-sectional study by Nankabirwa et al. [49] indicated that up to 39.9% of malaria cases in children under five years old are undiagnosed in Uganda. The under-reporting of malaria cases in Europe was estimated to vary between 20% to 59% by Legros and Danis [51]. This estimate is consistent with the 33.33% rate estimated for the Netherlands in 1996 by Van Hest et al. [52]. Overall, there are limited studies on the underestimation of malaria cases in the literature. To our knowledge only two studies have looked at the underestimation of malaria cases in Africa [48, 49]. To better understand the magnitude of malaria epidemic in Africa, it is essential to understand the degree of underestimation of cases and deaths.

In this study, we used a deterministic mathematical model to study the dynamics of malaria in Cameroon. Our model incorporates the human and mosquito population, and a seasonal mosquito biting rate. Using a Bayesian inference framework, we fitted our model to the reported cases of malaria in each of the ten regions of Cameroon, from January 2019 to December 2021, from which we estimated the underestimation of malaria cases in each region, and other key epidemiological parameters. In addition, through Hierarchical Clustering on Principal Components analysis (HCPC), we clustered the ten regions based on the quantified underestimation rates and other regional characteristics.

## Methods

### Mathematical model

We used a deterministic compartmental model to study the dynamics of malaria in the ten geographical regions of Cameroon, following the framework of [10, 33–36]. Our model considers the human and mosquito population in each region, dividing the human population into four compartments: susceptible (*S*_*h*_), exposed (*E*_*h*_), infectious (*I*_*h*_), and recovered (*R*_*h*_), and the mosquito population into two compartments: susceptible (*S*_*v*_) and infected (*I*_*v*_) (see Fig 1). Although many of the malaria models in the literature explicitly consider the population of mosquitoes in the exposed stage of the disease [35, 36], these mosquitoes are included in the infected compartment (*I*_*v*_) in our model.

**Fig 1.**
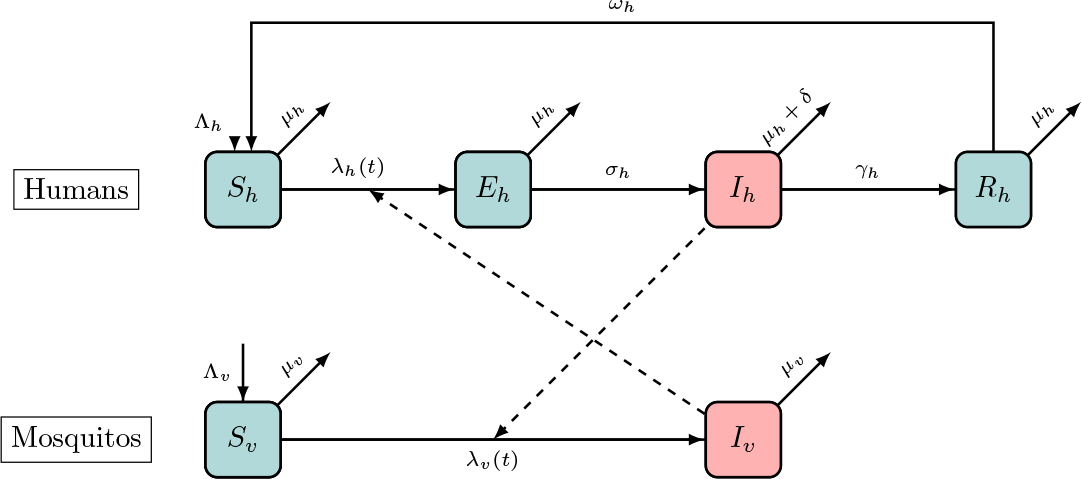
Schematic illustration of the compartmental model. An illustration of our model incorporating human and mosquito populations. The human population is divided into susceptible (*S*_*h*_), exposed (*E*_*h*_), infectious (*I*_*h*_), and recovered (*R*_*h*_) populations, while the mosquito population is divided into susceptible (*S*_*v*_) and infected (*I*_*v*_) mosquitoes. Solid black arrows show the transition of humans and mosquitoes through the different stages of malaria at the rate beside the arrows. Dashed black arrows show malaria transmission from humans to mosquitoes and vice versa.

In our model, a susceptible human becomes infected with malaria at a probability *β*_*h*_ following a bite from an infected mosquito. Similarly, a susceptible mosquito becomes infected with malaria at a probability *β*_*v*_ upon biting an infected human. After a successful bite and malaria transmission from a mosquito to a human, the malaria parasite in this individual undergoes an incubation period during which the infected humans cannot transmit the parasite to susceptible mosquitoes [37]. The incubation period lasts for a couple of days depending on the initial dose of the *plasmodium* parasite injected into the human by the mosquito [37]. This period is captured by the exposed compartment (*E*_*h*_) in our model. After this period, the infected individual becomes infectious and can transmit the parasite to susceptible mosquitoes. We assumed a constant influx of humans into each regions through birth and/or immigration at the rate Λ_*h*_, and similarly for mosquitoes at the rate Λ_*v*_. We did not explicitly model mosquito habitat and assumed that all the mosquitoes considered are the female *Anopheles* mosquitoes that can be infected with and transmit malaria [38, 39]. In addition to the natural human death rate *μ*_*h*_, we considered malaria-induced deaths in the human population at rate *δ*. To capture the seasonality of the malaria epidemic in each region, we assumed a periodic change in the biting rate of mosquitoes. We also incorporated the waning of infection-acquired temporary immunity in humans at rate *ω*_*h*_, which we assumed varies between the regions. The differential equations of the model are given by

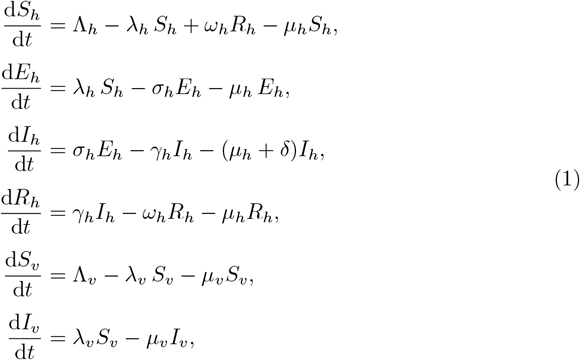

where *λ*_*h*_ ≡ *λ*_*h*_(*t*) and *λ*_*v*_ ≡ *λ*_*v*_(*t*) are the time-dependent forces of infection for the human and mosquito populations, respectively, given by

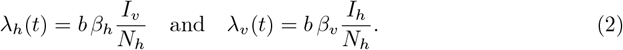

In Eq 2, *N*_*h*_ denotes the human population size, *b* denotes the biting rate of one mosquito per unit time, and *β*_*h*_ (*β*_*v*_) denotes the probability of malaria transmission to human (to a mosquito) upon one infectious bite. We assumed that one mosquito bites the same human only once during its lifetime.

To account for the seasonality in the biting rate of mosquitoes, we used a periodic function for *b* ≡ *b*(*t*), given by

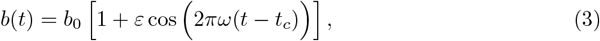

where *b*_0_ is the average biting rate of mosquitoes, *ε* is the degree of variation around the average biting rate, 1*/ω* is the period for seasonal malaria epidemic and *t*_*c*_ is the phase shift in the periodicity. All model parameters, their descriptions, and values are presented in Table 1.

**Table 1.** Descriptions and values for model variables and parameters.

| Variable | Description | Initial value | Source |
| --- | --- | --- | --- |
| $S_h$ | Susceptible human population | $S_h(0) = N_h(0) - (E_h(0) + I_h(0))$ | Assumed |
| $E_h$ | Exposed human population | See S2 File | Fitted |
| $I_h$ | Infectious human population | See S2 File | Fitted |
| $R_h$ | Recovered human population | $R_h(0) = 0$ | Assumed |
| $N_h$ | Total human population | See S2 File | [40] |
| $S_v$ | Susceptible mosquito population | $S_v(0) = 5,000$ | Assumed |
| $I_v$ | Infected mosquito population | See S2 File | Fitted |
| Parameter | Description | Value | Source |
| $\lambda_h(t)$ | Force of infection for human | See Eq 2 | - |
| $\lambda_v(t)$ | Force of transmission for mosquitoes | See Eq 2 | - |
| $b(t)$ | Time-dependent mosquito biting rate | See Eq 3 | - |
| $\Lambda_h$ | Human birth/immigration rate | See S2 File | [40] |
| $\Lambda_v$ | Mosquito birth/immigration rate | See S2 File | Fitted |
| $\beta_h$ | Probability of malaria transmission to human per bite | 0.022 | [41] |
| $\beta_v$ | Probability of malaria transmission to mosquito per bite | 0.48 | [41] |
| $\omega_h$ | Region-specific waning rate of malaria natural immunity in human | See S2 File | Fitted |
| $\mu_h$ | Human death rate | $0.0014 \text{ month}^{-1}$ | [42] |
| $\mu_v$ | Mosquito death rate | $0.033 \text{ day}^{-1}$ | [41] |
| $\sigma_h$ | Disease progression rate from exposed to infected for human | $0.1 \text{ day}^{-1}$ | [41] |
| $\gamma_h$ | Human recovery rate | $0.0035 \text{ day}^{-1}$ | [41] |
| $\delta$ | Human death rate due to malaria | See S2 File | Fitted |
| $p$ | Fraction of undetected malaria cases | See S2 File | Fitted |
| $b_0$ | Average biting rate of a mosquito | See S2 File | Fitted |
| $\varepsilon$ | Degree of variation around the average biting rate of mosquitoes | See S2 File | Fitted |
| $\omega$ | Frequency of seasonality in mosquito biting rate | See S2 File | Fitted |
| $t_c$ | Phase shift in the periodicity of mosquito biting rate | See S2 File | Fitted |

Using the next-generation matrix approach [43], we derived the time-dependent effective reproduction number ℛ_*e*_(*t*) of malaria for our model. Details of the derivation and the ℛ_*e*_(*t*) computed for each region using the median estimated parameters are presented in the S1 File.

### Data

We considered the monthly reported malaria cases in the ten geographical regions of Cameroon: Adamawa, Central, East, Far North, Littoral, North, Northwest, West, South, and Southwest, from January 2019 to December 2021, as obtained from DHIS2 [44]. The data is stratified into two age groups: *<* 5 years old and ≥ 5 years old (see Fig 2). However, in this study, we combined the data for the two age groups to obtain the total estimated cases of malaria for each region, which is used in our model calibration.

**Fig 2.**
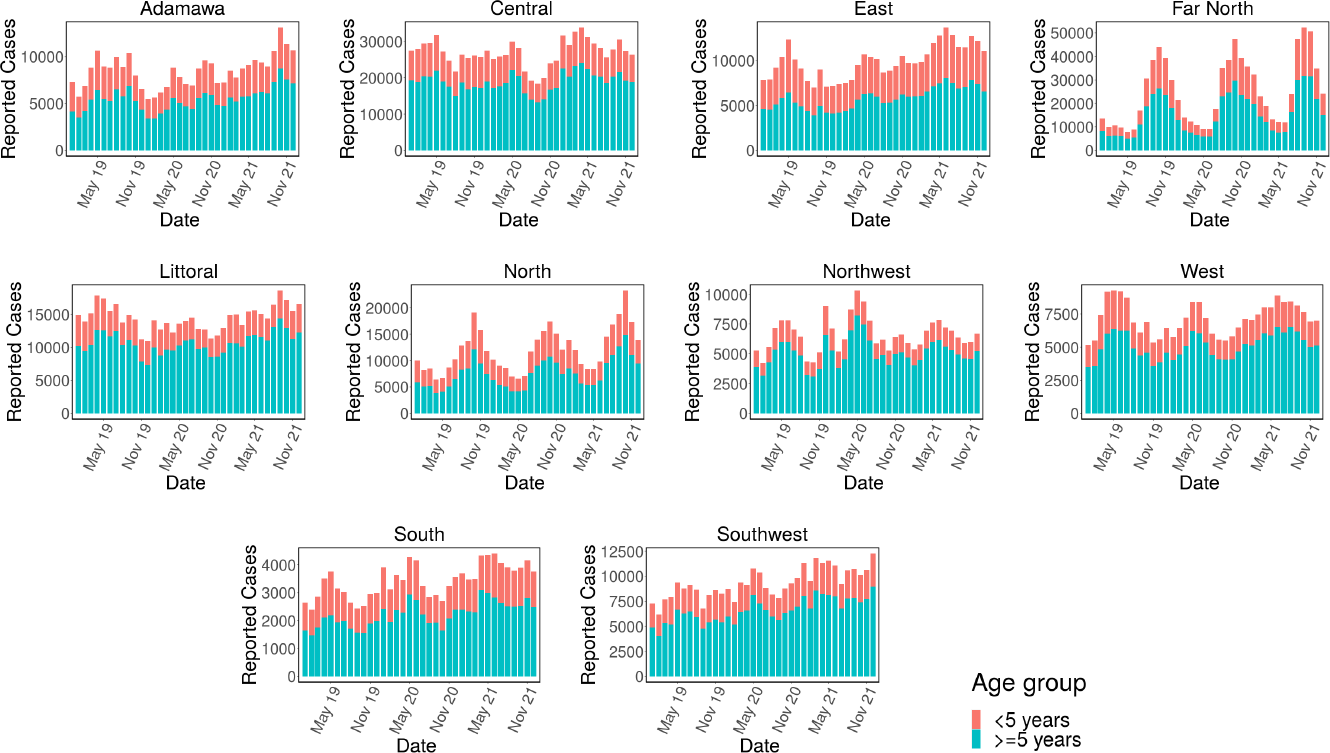
Monthly reported cases of malaria. The monthly reported cases of malaria in different geographical regions of Cameroon from January 2019 to December 2021, for different age groups, obtained from DHIS2 [44]. Red bars represent the reported cases for *<* 5 years age-group, and the green bars is for the ≥ 5 years age-group.

Fig 2 shows the bar plots of the monthly reported cases of malaria for each region for the *<* 5 years old (red bars) and those ≥ 5 years old (green bars). The seasonality in the total cases of malaria in each region can easily be seen. Many of the regions have only one (usually large) outbreak of malaria each year. However, the Northwest region has multiple smaller outbreaks yearly.

### Bayesian inference

We fitted our model in Eq (1) to the monthly reported cases of malaria in each region independently using a Bayesian inference framework and the RStan package in R version 3.6.3 [45]. Although the data are stratified by age groups when presented in Fig 2, we used the total reported cases (without age stratification) in our fit. In our calibration, we defined the likelihood function for each region as

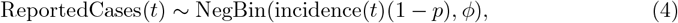

where NegBin(·) is the negative binomial distribution, ReportedCases(*t*) is the total monthly reported cases of malaria for the region obtained from the data in Fig 2, incidence(*t*) is the monthly incidence of malaria computed using our model Eq 1 (it is computed as the number of exposed individuals transitioning to the infectious compartment each month), *p* is the fraction of cases undetected (i.e., 1 − *p* is the fraction of malaria cases that was detected), and *ϕ* is the dispersal parameter.

The Bayesian inference framework gives us the flexibility to incorporate our prior knowledge into the model parameters and the ability to evaluate probabilistic statements of the data based on the model. We implemented uninformative priors when fitting our model to the case data. To ensure that our model is coded accurately in the Stan and also for validation, we generated synthetic case data using our model with known parameter values. We then calibrated our model to the synthetic dataset, and inspected the resulting posterior distributions for biases and coverage of the true parameter values used to generate the data. We also checked for the identifiability of the parameters. We used the adaptive Hamiltonian Monte Carlo method No-U-Turn sampling (NUTS) as implemented in RStan with 5,000 iterations and four chains for our model calibrations. From our fit, for each region, we estimated the fraction of underestimated cases of malaria (*p*), the average biting rate of a mosquito (*b*_0_), the malaria-induced death rate for humans (*δ*), and other important model parameters (see Results Section). These parameters are used to compute the time-dependent effective reproduction number ℛ_*e*_ of malaria for each region (See S1 File).

## Results

### Data fitting

We calibrated our model in Eq (1) to the monthly reported cases of malaria in each of the ten regions of Cameroon (see Fig 2) independently using a Bayesian inference framework. The results of our inference showing the monthly reported malaria cases (black dots), together with the median predicted cases, obtained from our model fit (solid lines), are presented in Fig 3. The narrower (darker) bands are 50% credible intervals (CrI), while the wider (lighter) bands are 90% CrI.

**Fig 3.**
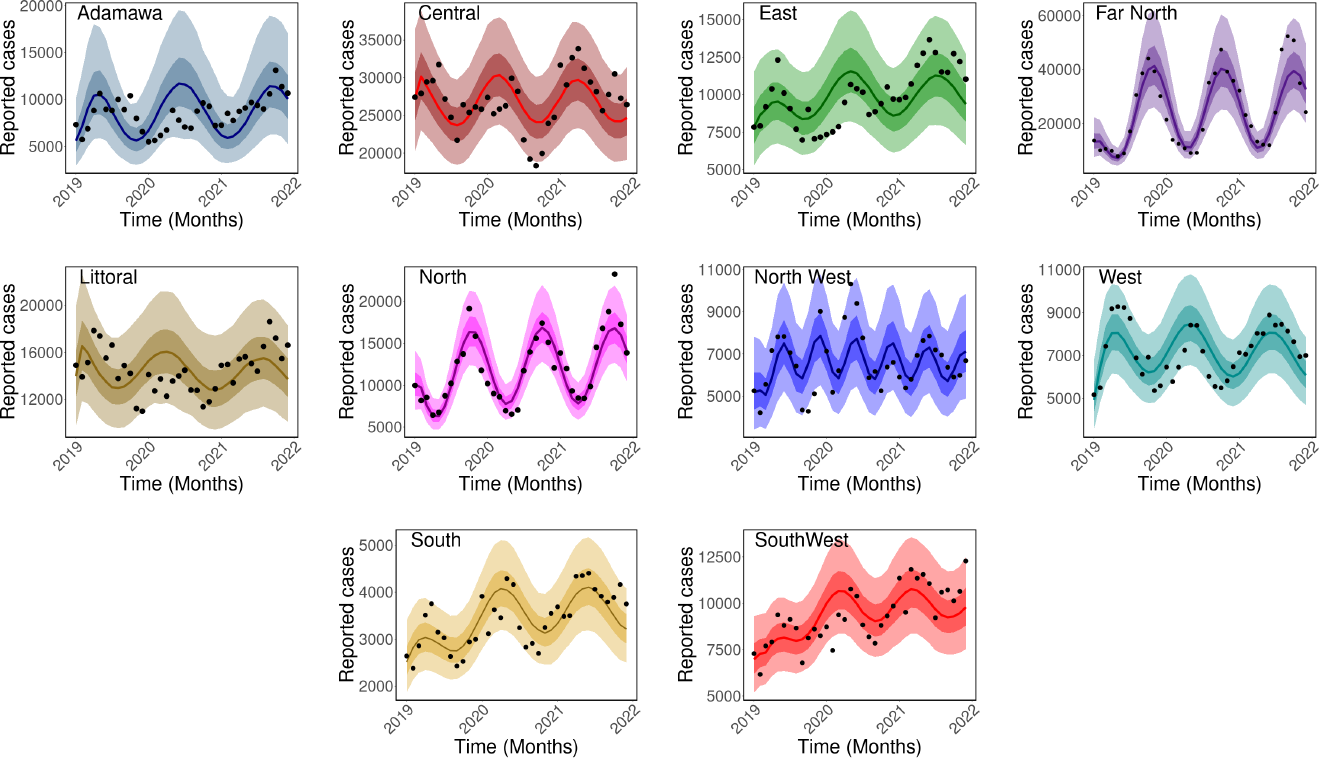
Observed and estimated monthly malaria cases. The monthly reported malaria cases (black dots) and median predicted cases (solid lines) for each region of Cameroon. The darker bands are the 50% CrI, while the lighter bands are the 90% CrI.

### Quantifying the underestimation of malaria cases

From our calibration, we estimated the percentage of underestimated cases of malaria in each region, which varies significantly between the regions (Fig 4). The smallest percentage was estimated for the East region with a median of 14% (90% CrI: 1% - 38%), while the largest was estimated for the Northwest region, 70% (90% CrI: 40% - 83%). Following the Northwest region is the Littoral and West regions, with similar rates, 67% (90% CrI: 39% - 80%) and 67% (90% CrI: 47% - 76%), and then Far North, 48% (90% CrI: 15% - 68%) and North, 47% (90% CrI: 13% - 68%).

**Fig 4.**
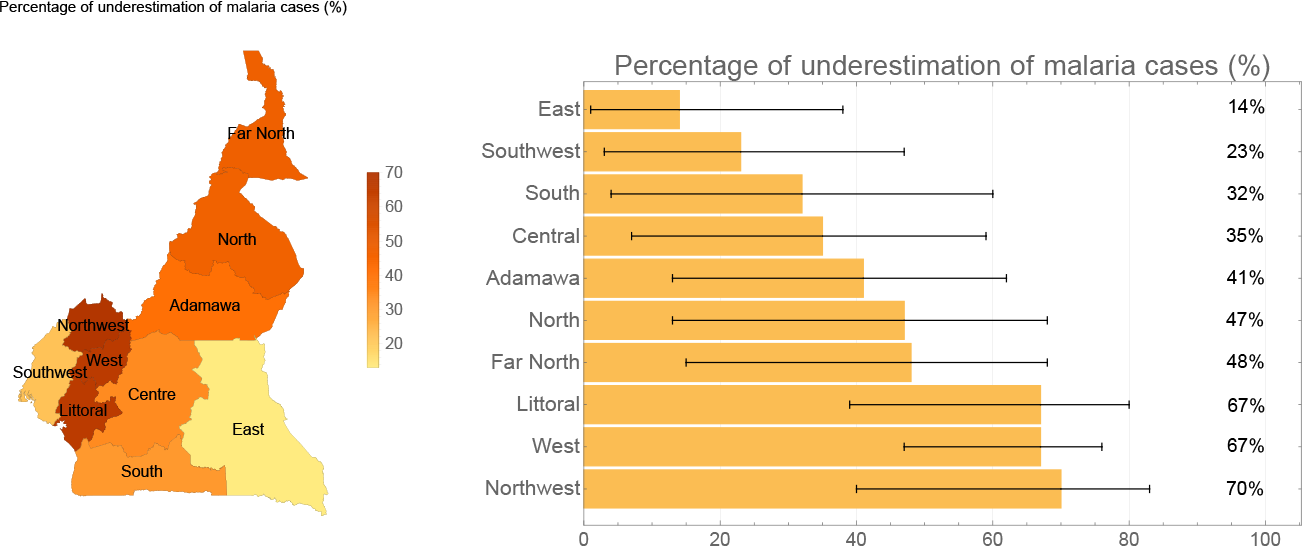
Percentage of malaria cases underestimation. The percentage of underestimation of malaria cases estimated for each region. Left panel: map visualization with the median underestimation percentage indicated with color intensity: high (reddish) and low (yellowish). Right panel: barplots showing the percentage of underestimation for each region with 90% CrI (in ascending order).

We present five key malaria parameters estimated from our fit in Fig 5 and Fig 6: the mean duration of natural immunity (1*/ω*_*h*_) in months, malaria-induced per capita death rate (*δ*), the period of seasonal mosquito biting rate (1*/ω*) in months, the baseline (or average) mosquito biting rate (*b*_0_) per day, and the degree of variation around baseline biting rate (*ε*) in percentage.

**Fig 5.**
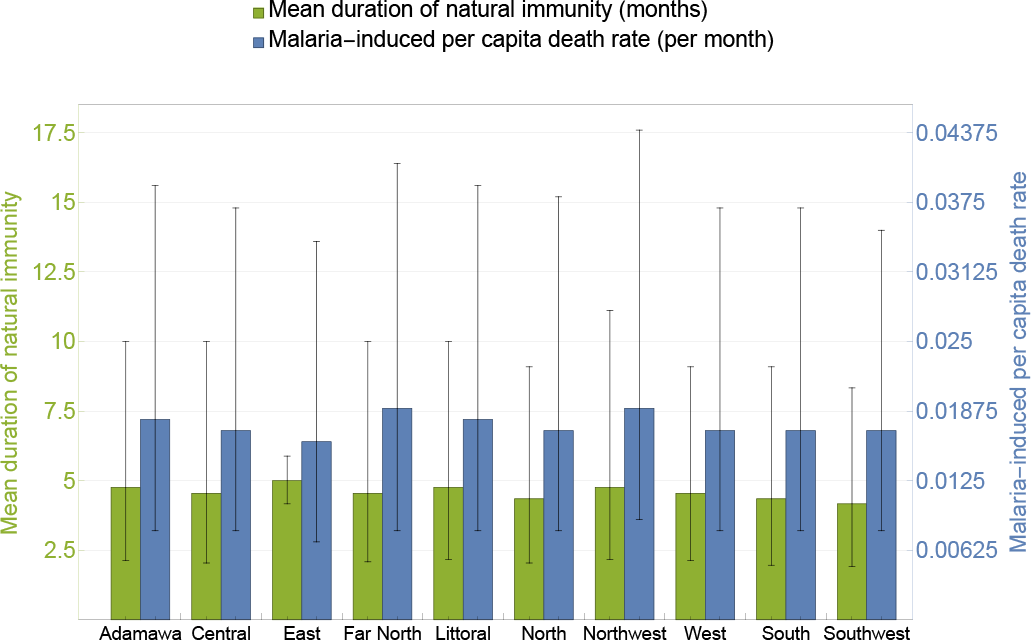
Estimated region-specific malaria parameters: mean duration of natural immunity in months and malaria-induced per capita death rate per month. The bars are the median estimated values and error bars are for the 90% CrI.

**Fig 6.**
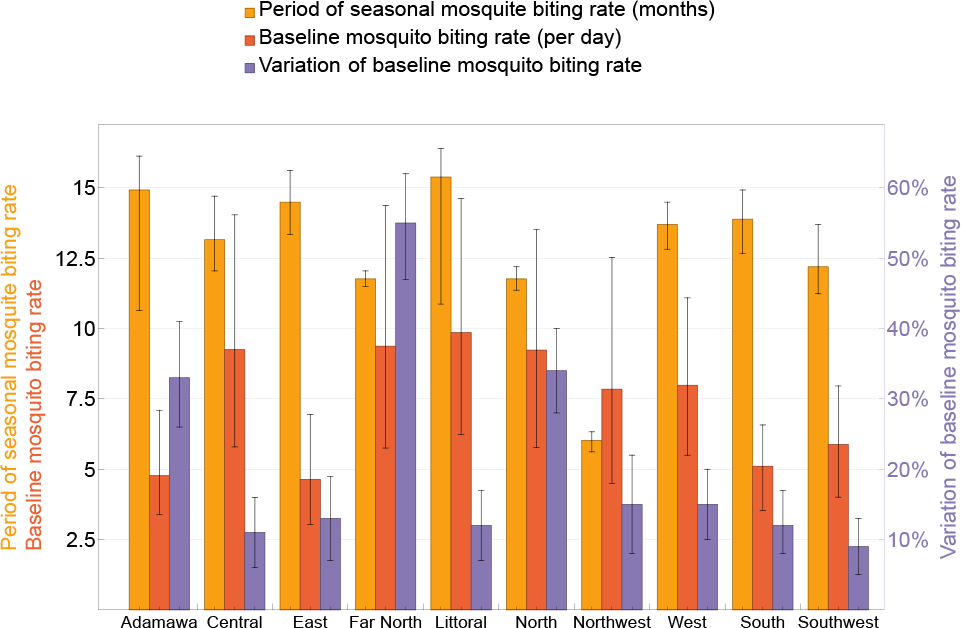
Estimated region-specific malaria parameters: period of oscillation of mosquito biting rate in months, baseline mosquito biting rate per day and variation of baseline mosquito biting rate in percentage. The bars represent the median estimated values and error bars are for the 90% CrI.

The estimated malaria-induced death rate is similar for the regions ranging from a median of 0.016/month (90% CrI: 0.007-0.034) for the East region to 0.019/month for Far North (90% CrI: 0.008-0.041) and Northwest (90% CrI: 0.009-0.044). The mean duration of natural immunity is also consistent among the regions (see Fig 5). The East region has a natural immunity duration of 5 months (90% CrI: 4.17-5.88) and the Southwest region has the shortest duration of 4.17 months (90% CrI: 1.92-8.33). It is important to note that the East region has the highest estimation rates of malaria cases while it has the lowest malaria-induced death rates and longest natural immunity duration.

We estimated that the mosquito biting rate for the Northwest region oscillated with a period of 6.02 months (90% CrI: 5.62-6.33), while those of the other regions oscillated with a period of at least 12 months, with the biting rate of Littoral having the largest periodicity of 15.38 months (90% CrI: 10.87-16.39). The low periodicity of mosquito biting rate in the Northwest region may have contributed to the high underestimation of malaria cases estimated for this region. Most of the regions have an estimated mosquito biting rate above 5 bite/day, with the highest estimated for the Littoral region with a median of 9.86 bites/day (90% CrI: 6.24-14.61). Two regions have biting rates below 5 bites/day, and they are the Adamawa region with a median of 4.78 bites/day (90% CrI: 3.39-7.09), and the East region with 4.64 bites/day (90% CrI: 3.03-6.94). The degree of variation around baseline mosquito biting rate was estimated to be mostly between 9% and 35%, with Far North having the only exceptionally large variation of 55% (90% CrI: 47%-62%), and Southwest having the least variation of 9% (90% CrI: 5%-13%).

### Clustering of regions

We used the estimated malaria underestimation rates, the population sizes, total numbers of healthcare human resources per 1,000, and total numbers of healthcare facilities per 1,000 to cluster the ten Cameroon regions through Hierarchical Clustering on Principal Components (HCPC).

HCPC resulted in four major clusters as shown in Fig 7A, indicated by different colors. Cluster 1 includes Aadmawa alone, cluster 2 includes the South, East and Southwest regions, cluster 3 contains the West and Northwest regions, and cluster 4 has Littoral, Far North, North and Central. Among the four factors used for this clustering, the fraction of undetected cases (*p*=0.018), number of healthcare human resources (*p*=0.0043) and population size (*p*=0.0074) are significantly different across clusters from ANOVA test, as shown in Fig 7B. However, these clusters do not differ in number of healthcare facilities (*p*=0.19).

**Fig 7.**
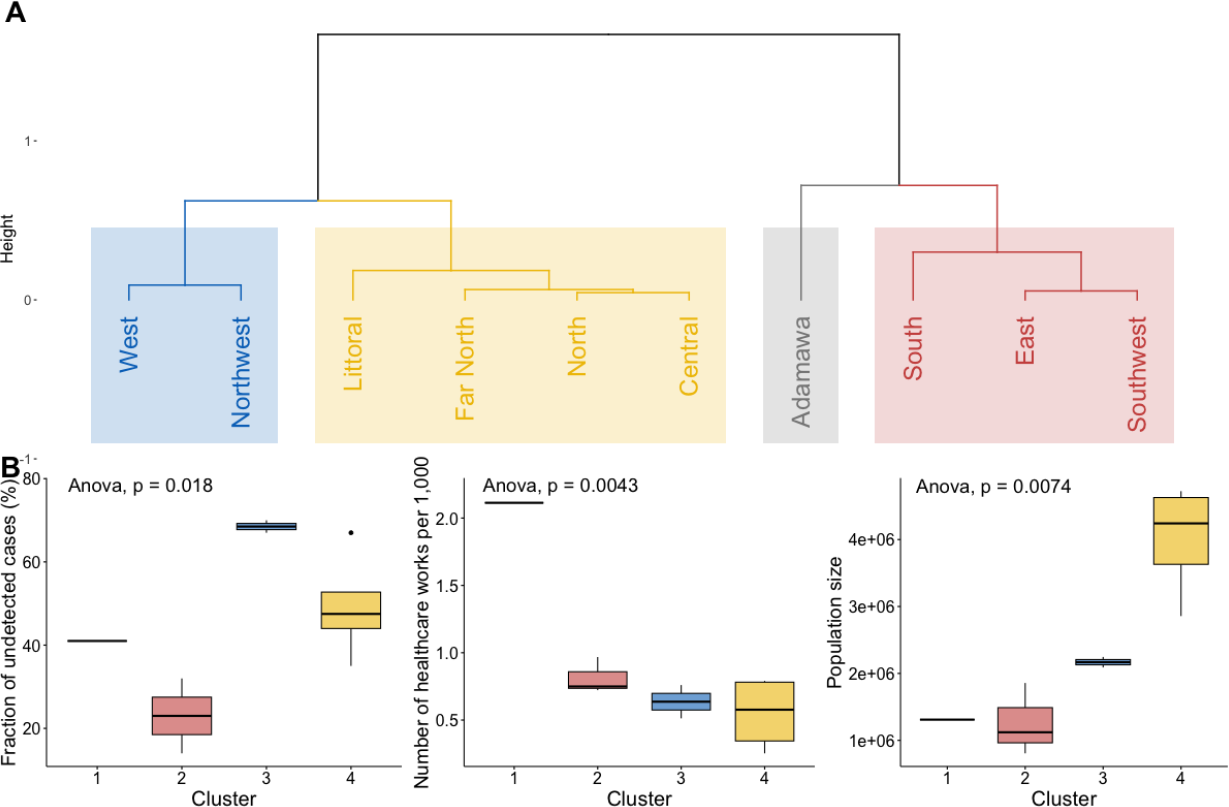
Results from Hierarchical Clustering on Principal Components (HCPC). A: Dendrogram showing the clustering of the Cameroon regions. B: Significant variables characterizing the clusters, with p-values from ANOVA test.

## Discussion

We have used a deterministic mathematical model to quantify the underestimation of malaria cases in ten regions of Cameroon. The research aimed to estimate essential epidemiological parameters and reveal the extent of underestimation in malaria dynamics across the country. Our findings showed significant variations in the degree of underestimation, ranging from 14% in the East region to a substantial 70% in the Northwest region. We observe from our model fit that the Northwest region has the largest number of smaller epidemics that occurred during our study period of January 2019 to December 2021, among the regions. Although these epidemics do not result in high number of malaria cases like those reported in other regions (Adamawa, Central, East, Far North, Littoral North, and Southwest), the frequency of the epidemics may have resulted in the high percentage of underestimation obtained for this region.

Furthermore, the research indicated a consistent malaria-induced death rate across Cameroon, with the East region reporting the lowest rate at 0.016/month, while the Far North and Northwest regions recorded the highest at 0.019/month. The mean duration of natural immunity displayed uniformity among regions, with all regions showing a duration of less than five months. Notably, the East region had a relatively longer natural immunity duration of 5 months, while the Southwest region had the shortest at 4.17 months. Moreover, the study revealed that only the Northwest region exhibited a minor period of oscillation for the mosquito biting rate (6.02 months), while all other regions demonstrated oscillation periods greater than or equal to 12 months. The Littoral region exhibited the longest periodicity at 15.38 months, indicating substantial variation in mosquito activity across Cameroon.

Regarding mosquito biting rates, most regions had estimated rates exceeding 5 bites per day, with the Littoral region topping the list at a median of 9.86 bites per day. In contrast, only two regions reported biting rates below 5 bites per day, specifically the Adamawa region (4.78 bites/day) and the East region (4.64 bites/day).

To gain insights into regional disparities, we conducted Hierarchical Clustering on Principal Components analysis, considering undetected fractions, total population size, total healthcare human resources per 1000, and total healthcare facilities per 1000. This analysis resulted in four major clusters: Cluster 1 consisted of Adamawa alone, Cluster 2 included the South, East, and Southwest regions, Cluster 3 contained the West and Northwest regions, and Cluster 4 comprised the Littoral, Far North, North, and Central regions. Significantly, the fraction of undetected cases (p=0.018), the number of healthcare human resources (p=0.0043), and population size (p=0.0074) exhibited notable differences across these clusters, as determined by the ANOVA test. However, the number of healthcare facilities did not display significant variation (p=0.19) among the clusters.

In the current academic literature, there have been a limited number of efforts to assess the hidden impact of malaria in both malaria-prone and non-malaria-prone regions worldwide. For instance, in malaria-prone areas, Chourasia et al. [46] estimated the burden of asymptomatic malaria among a tribal population in a forested village in central India. They used peripheral blood smears from 134 and 159 individuals in the village during 2013 and 2014. The authors found that the prevalence rates of asymptomatic malaria were 20% and 22.8% for the two years, respectively. Yadav et al. [47] conducted a retrospective exploration of surveillance data and health records from major public and private health facilities in Ahmedabad city, India. They observed that the detected malaria incidence (37,431 cases) was nine times higher than what was officially reported (4,119 cases), meaning that only 11% of the total malaria cases were reported. In the Ifanadiana District in Madagascar, passive surveillance was estimated to have reported 1 in every 5 malaria cases among all individuals (20%), and 1 in every 3 cases among children under five (33.33%) from 2014 to 2017 [48]. Nankabirwa et al. [49] conducted a cross-sectional study that revealed 39.9% of children under five years old in Uganda are under-diagnosed for malaria. Breman [50] observed that less than 20% of malaria incidence cases in sub-Saharan Africa reach the formal health system. In non-malaria-prone regions of the world, Legros and Danis [51] estimated the under-reported rate of malaria in Europe varies from 20% to 59%. This is consistent with Van Hest et al. [52] estimating that approximately one-third (33.33%) of cases in the Netherlands in 1996 went unreported and Cartcath et al. estimating that 34% of cases in England went unreported from 2003-2004. Based on these studies, there isn’t much disparity in malaria underestimation between malaria-prone regions and non-malaria-prone regions. Our study reveals that the mean undetected malaria cases across Cameroon is 44.4% (S.D. 19.24%), which aligns with the undetected values in the studies above.

The studies mentioned were identified through searches in two major scholarly electronic databases in the biomedical field, namely, MEDLINE via its openly accessible interface, PubMed, and Scopus. To ensure we captured all relevant studies that hadn’t been indexed, we also conducted searches on Google Scholar. The following search strings were utilized: (“malaria” OR “malarial fever”) AND( (“Underestimate” OR “Reporting gap” OR “Hidden cases” OR “Under-declared” OR “Unrecorded” OR “Missed reports” OR “Under-ascertainment” OR “Under ascertainment” OR “Reporting bias” OR “Undercount” OR “under reporting” OR “underreporting” OR “underascertainment”) OR (“misdiagnosis” OR “Missed diagnosis” OR “False negative” OR “Incorrect diagnosis” OR “Underdiagnosis” OR “Failed detection” OR “Diagnostic error” OR “Diagnostic bias” OR “Undetected” OR “Under-assessment”) OR (“Under-assess” OR “Under-rate” OR “underrate” OR “Under-ascertain” OR “Underestimate bias” OR “Inaccurate assessment” OR “under-estimation” OR “Underestimation”)OR (“Non-reporting” OR “Unreported “OR “Under-declared “OR “Underdeclared” OR “Missed notification” OR “Notification deficit” OR “Silent reporting” OR “Notification bias” OR “Notification-bias” OR “Underdocumented” OR “Under-registered” OR “Underestimated”))AND (“mathematical model” OR “statistical model” OR “model-based study” OR “modeling study” OR “model*”).

We found that in the existing scholarly literature, there is no mechanistic model for estimating the underestimated proportions of malaria. As a result, our model is the first mechanistic model designed for this purpose. The observation that underestimated fractions and the number of healthcare personnel are variables defining clusters is quite significant and raises some important points for investigation. The significance of underestimated fractions within clusters alongside a low number of healthcare personnel suggests a potential correlation. It may imply that regions with fewer healthcare professionals are more likely to under-estimate cases.

Our study has several limitations that should be acknowledged, including the limited variables chosen for clustering analysis and the omission of an assessment of the factors responsible for underestimation. Future studies should explore the impact of a wide array of socio-economic, climatic, epidemiological, health system-related, demographic, and clinical public health variables on the underestimation of malaria. Further research is warranted to investigate the impact of climatic factors, such as rainfall and temperature on the dynamics of malaria across Cameroon. Seasonal variations in these factors can affect mosquito populations and biting habits, which, in turn, may influence the spread of malaria

In conclusion, factors like self-administered treatment, the availability of healthcare facilities, the healthcare workforce, the presence of asymptomatic malaria cases, the capacity of medical laboratories, and the quality of sanitation infrastructure all have the potential to influence people’s inclination to reveal their malaria status or to seek medical help. Recognizing these variations is essential for designing customized and comprehensive strategies that can efficiently tackle underestimation, ultimately resulting in more significant and enduring improvements in clinical public health. The notably low reported rates in Cameroon indicate a distinct necessity for the enhancement of reporting and surveillance systems.

## Supporting information

S1 File: Effective reproduction numbers.

## Data Availability

The monthly reported cases of malaria in Cameroon were obtained from DHIS2 (https://dhis2.org), and are available within the article and its Supplementary material as well as on the link below: https://acadic.org/africa-in-data/. DHIS2 is an open-source, web-based software platform for data collection, management, and analysis in all government hospitals and public health centers in the countries where aggregation of health data from both government and non-government hospitals is collected. The Bayesian inference code used for our model calibration is available on https://github.com/iyaniwura/Malaria_Model_Fit_Code

https://github.com/iyaniwura/Malaria_Model_Fit_Code

## Supporting information

**S1 File. Effective reproduction numbers.**

**S2 File. Supplementary tables**.

## Data availability

The monthly reported cases of malaria in Cameroon were obtained from DHIS2 [44], and are available within the article and its Supplementary material. DHIS2 is an open source, web-based software platform for data collection, management, and analysis in all government hospitals in the countries where aggregation of health data from both government and non-government hospitals is collected. The Bayesian inference code used for our model calibration is available on GitHub at https://github.com/iyaniwura/Malaria_Model_Fit_Code.

## Acknowledgments

JDK acknowledges support from New Frontier in Research Fund-Exploratory (Grant No. NFRFE-2021-00879) and NSERC Discovery Grant (Grant No. RGPIN-2022-04559). Portions of this work were performed at the Los Alamos National Laboratory under the auspices of the US Department of Energy contract 89233218CNA000001 and supported by NIH grant R01-OD011095.

