## Supplementary material for "Regional variation and epidemiological insights in malaria underestimation in Cameroon": S1 File: Effective reproduction numbers.

We computed the effective reproduction number of malaria for our model using the next-generation matrix approach [1]. To do this, we construct the matrix $\mathcal{F}$, which describes new infections to the system, and matrix $\mathcal{V}$, which describes all other transitions within the system, using the infected compartments $E_h, I_h$ and $I_v$.

$$\mathcal{F} = \begin{bmatrix} \lambda_h S_h \\ 0 \\ \lambda_v S_v \end{bmatrix}, \quad \mathcal{V} = \begin{bmatrix} \sigma_h E_h + \mu_h E_h \\ -\sigma_h E_h + \gamma_h I_h + (\mu_h + \delta) I_h \\ \mu_v I_v \end{bmatrix}.$$

Taking the partial derivatives of these matrices and evaluating them at the $S_h(t), S_v(t)$ and $N_h(t)$ yields

$$F = \begin{bmatrix} 0 & 0 & b(t)\beta_h \frac{S_h(t)}{N_h(t)} \\ 0 & 0 & 0 \\ 0 & b(t)\beta_v \frac{S_v(t)}{N_h(t)} & 0 \end{bmatrix}, \quad V = \begin{bmatrix} \sigma_h + \mu_h & 0 & 0 \\ -\sigma_h & \gamma_h + \mu_h + \delta & 0 \\ 0 & 0 & \mu_v \end{bmatrix},$$

and

$$FV^{-1} = \begin{bmatrix} 0 & 0 & b(t)\beta_h \frac{S_h(t)}{N_h(t)} \frac{1}{\mu_v} \\ 0 & 0 & 0 \\ b(t)\beta_v \frac{S_v(t)}{N_h(t)} \frac{\sigma_h}{(\sigma_h+\mu_h)(\gamma_h+\mu_h+\delta)} & b(t)\beta_v \frac{S_v(t)}{N_h(t)} \frac{1}{\gamma_h+\mu_h+\delta} & 0 \end{bmatrix}.$$

Therefore, the effective reproduction number for malaria is given by

$$\mathcal{R}_e(t) = \sqrt{K_{hv}(t) K_{vh}(t)}, \tag{1}$$

with

$$\begin{aligned} K_{hv}(t) &= b(t)\beta_v \frac{S_v(t)}{N_h(t)} \left[ \frac{\sigma_h}{(\sigma_h + \mu_h)(\gamma_h + \mu_h + \delta)} \right], \\ K_{vh}(t) &= b(t)\beta_h \frac{S_h(t)}{N_h(t)} \frac{1}{\mu_v}. \end{aligned} \tag{2}$$

$K_{hv}(t)$ is the average number of infected mosquitoes one infectious human can produce during his/her entire infectious period at time $t$ and $K_{vh}(t)$ is average number of infected humans one infectious mosquito can generate during its whole period of infectiousness at time $t$. $\mathcal{R}_e(t)$ is the geometric average of the two.

Using the median estimated parameters from our model fit, we plotted the time-dependent effective reproduction number $\mathcal{R}_e$ in Fig 1. After dropping from the initial high value, the effective reproduction number oscillates around 1 for all regions, with larger magnitudes for Far North, North, and Adamawa. This agrees with the large variations around baseline mosquito biting rates for these three regions. The periodicity of $\mathcal{R}_e$ also agrees with the results in the main text. As Northwest exhibits almost two cycles every year and others roughly one cycle each year.

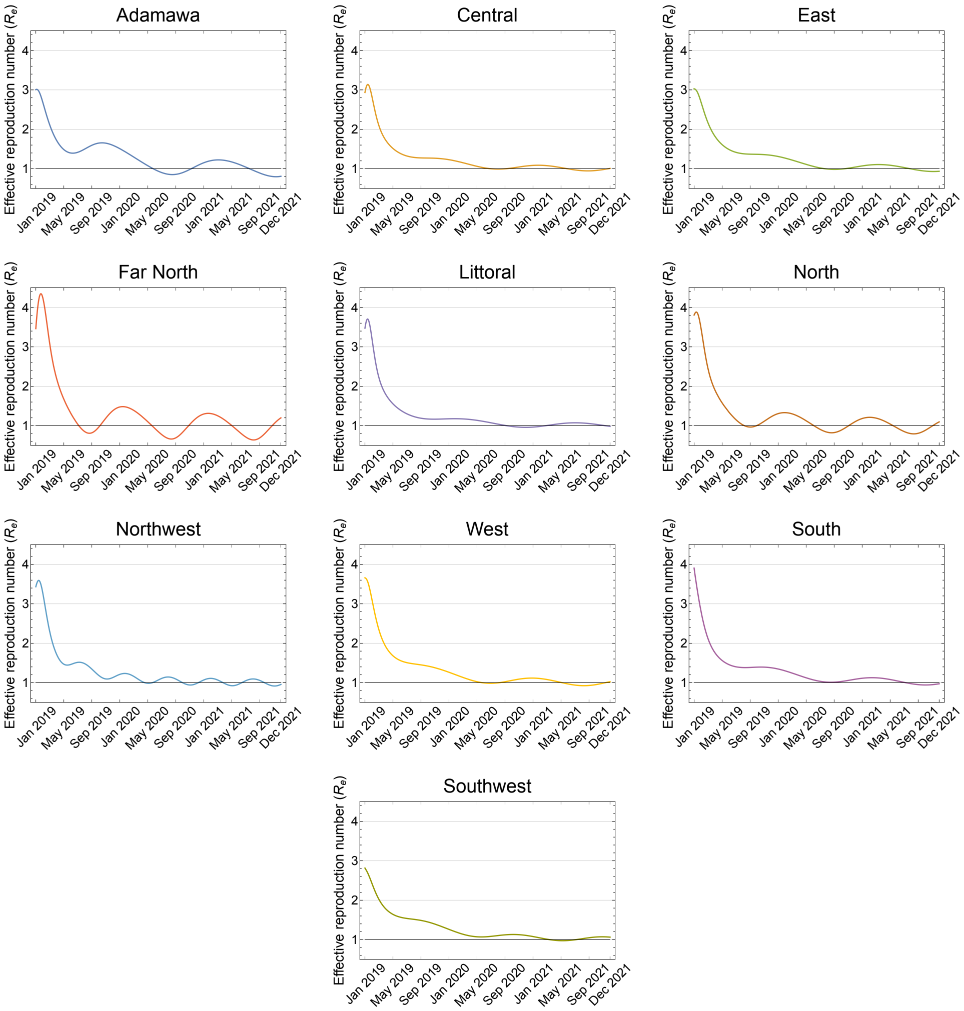


**Fig 1. Estimated region-specific effective reproduction number ($\mathcal{R}_e$).** The median estimated parameter values from our model fit were used to generate these results.
