## Supplementary material for "Regional variation and epidemiological insights in malaria underestimation in Cameroon": S2 File: Supplementary tables.

**Table 1. Estimated initial values and birth/immigration rate of mosquitoes with 90% credible interval.** The estimated initial infected humans ($E_h(0) + I_h(0)$), where $E_h(0)$ and $I_h(0)$ are the initial population of individuals in the exposed and infectious stages of malaria. $I_v(0)$ is the initial population of infected mosquitoes, and $\Lambda_v$ is the birth/immigration rate of mosquitoes.

| **Region** | $E_h(0) + I_h(0)$ | $I_v(0)$ | $\Lambda_v$ |
|---|---|---|---|
| Adamawa | 510 (95.6, 2,502) | 4,893 (2,771, 8,548) | 6,568 (3,958, 11,254) |
| Central | 506 (100, 2,444) | 12,593 (6,157, 28,822) | 8,448 (4,389, 19,498) |
| East | 524 (97.7, 2,522) | 5,373 (2,693, 10,959) | 5,333 (3,145, 10,182) |
| Far North | 503 (102, 2,372) | 6,866 (2,883, 16,881) | 9,872 (4,468, 24,751) |
| Littoral | 503 (101, 2,388) | 11,441 (4,750, 25,938) | 8,372 (3,491, 19,359) |
| North | 520 (96.5, 2,602) | 5,777 (2,487, 14,442) | 4,641 (2,165, 11,584) |
| Northwest | 517 (99.6, 2,596) | 6,643 (1,989, 19,422) | 5,759 ( 1,965, 16,370) |
| West | 515 (98.8, 2,601) | 5,074 (2,274, 10,559) | 5,519 (2.702, 11,025) |
| South | 843 (119, 4,172) | 1,541 (47.4, 4,452) | 2,200 (1,353, 5,049) |
| Southwest | 571 (101, 2,978) | 4,305 (2,303, 9,440) | 4,470 (2,706, 9,182) |

**Table 2. Estimated malaria cases underestimation fraction ($p$), average biting rate of mosquitoes per month ($b_0$), malaria-induced death rate per month ($\delta$), and natural immunity waning rate per month ($\omega_h$) with 90% credible interval.**

| **Region** | $p$ | $b_0$ | $\delta$ | $w_h$ |
|---|---|---|---|---|
| Adamawa | 0.41 (0.13, 0.62) | 143.3 (101.7, 212.8) | 0.018 (0.008, 0.039) | 0.21 (0.10, 0.47) |
| Central | 0.35 (0.07, 0.59) | 277.6 (173.8, 421.2) | 0.017 (0.008, 0.037) | 0.22 (0.10, 0.49) |
| East | 0.14 (0.01, 0.38) | 139.3 (90.9, 208.3) | 0.016 (0.007, 0.034) | 0.20 (0.17, 0.24) |
| Far North | 0.48 (0.15, 0.68) | 281.2 (172.6, 431.3) | 0.019 (0.008, 0.041) | 0.22 (0.10, 0.48) |
| Littoral | 0.67 (0.39, 0.80) | 295.7 (187.1, 438.4) | 0.018 (0.008, 0.039) | 0.21 (0.10, 0.46) |
| North | 0.47 (0.13, 0.68) | 277.0 (172.9, 405.4) | 0.017 (0.008, 0.038) | 0.23 (0.11, 0.49) |
| Northwest | 0.70 (0.40, 0.83) | 235.4 (134.9, 375.7) | 0.019 (0.009, 0.044) | 0.21 (0.09, 0.46) |
| West | 0.67 (0.47, 0.76) | 239.6 (164.9, 332.7) | 0.017 (0.008, 0.037) | 0.22 (0.11, 0.47) |
| South | 0.32 (0.04, 0.60) | 153.2 (106.2, 197.2) | 0.017 (0.008, 0.037) | 0.23 (0.11, 0.51) |
| Southwest | 0.23 (0.03, 0.47) | 176.4 (120.2, 238.8) | 0.017 (0.008, 0.035) | 0.24 (0.12, 0.52) |

**Table 3. Estimated degree of variation around the average biting rate of mosquitoes ($\varepsilon$), frequency of seasonality in mosquito biting rate ($\omega$), and phase shift in the periodicity of mosquito biting rate ($t_c$) with 90% credible interval.**

| Region | $\varepsilon$ | $w$ | $t_c$ |
|---|---|---|---|
| Adamawa | 0.33 (0.26, 0.41) | 0.067 (0.062, 0.094) | -2.53 (-2.70, -2.37) |
| Central | 0.11 (0.06, 0.16) | 0.076 (0.068, 0.083) | -0.61 (-2.04, 1.00) |
| East | 0.13 (0.07, 0.19) | 0.069 (0.064, 0.075) | -1.52 (-2.76, 0.084) |
| Far North | 0.55 (0.47, 0.62) | 0.085 (0.083, 0.087) | 1.94 (1.39, 2.57) |
| Littoral | 0.12 (0.07, 0.17) | 0.065 (0.061, 0.092) | 0.32 (-0.18, 0.85) |
| North | 0.34 (0.28, 0.40) | 0.085 (0.082, 0.088) | 2.28 (1.56, 3.05) |
| Northwest | 0.15 (0.08, 0.22) | 0.166 (0.158, 0.178) | 1.11 (0.10, 1.98) |
| West | 0.15 (0.10, 0.20) | 0.073 (0.069, 0.078) | -2.49 (-3.55, -1.07) |
| South | 0.12 (0.08, 0.17) | 0.072 (0.067, 0.079) | -1.25 (-2.34, 0.014) |
| Southwest | 0.09 (0.05, 0.13) | 0.082 (0.073, 0.089) | -2.00 (-2.81, -1.17) |

**Table 4. Human population size ($N_h$) and birth/immigration rate ($\Lambda_h$) for each region.** The population sizes and birth/immigration rates are based on the Cameroonian Ministry of Public Health estimate [1]. The birth/immigration rate for each region is the average monthly change in the population of the region between January 2019 and December 2021.

| Region | Population size (2019) | Human birth/immigration rate (per month) |
|---|---|---|
| Adamawa (Adamaoua) | 1,309,666 | 3,080 |
| Central (Center) | 4,723,372 | 10,327 |
| East (Est) | 1,120,567 | 2,171 |
| Far North (Extreme-Nord) | 4,595,669 | 11,885 |
| Littoral (Littoral) | 3,887,698 | 8,352 |
| North (Nord) | 2,856,872 | 8,806 |
| Northwest (Nord-Ouest) | 2,246,302 | 2,570 |
| West (Ouest) | 2,089,297 | 1,968 |
| South (Sud) | 805,741 | 1,067 |
| Southwest (Sud Ouest) | 1,857,169 | 3,294 |

**Table 5. Epidemiological factors considered for each region in our clustering analysis.**

| Region | Total number of healthcare human resources (2019) [2] | Total number of healthcare facilities (2023) |
|---|---|---|
| Adamawa (Adamaoua) | 2,768 | 136 |
| Central (Center) | 3,677 | 459 |
| East (Est) | 840 | 171 |
| Far North (Extreme-Nord) | 1,162 | 383 |
| Littoral (Littoral) | 3,073 | 200 |
| North (Nord) | 1,073 | 249 |
| Northwest (Nord-Ouest) | 1,152 | 244 |
| West (Ouest) | 1,588 | 415 |
| South (Sud) | 780 | 199 |
| Southwest (Sud Ouest) | 1,341 | 204 |
